# Global, National, and Regional Burden of Depressive Disorders in Menopause Women Aged 45--54 Years from 1990--2021

**DOI:** 10.1101/2025.04.26.25326495

**Authors:** Yufan Tang, Yanglu Hong, Shan Liu, Wei Zhou, Tian Gong, Yueshan Pang

**Affiliations:** Department of General Practice, Beijing Anzhen Nanchong Hospital of Capital Medical University & Nanchong Central Hospital, Sichuan, China

**Keywords:** Depression, Menopause, Epidemiology, Mental Disorders, Disease Burden

## Abstract

To analyze the trend changes in the burden of depressive disorders (DD) among menopausal women aged 45--54 years from 1990--2021 globally and to provide theoretical evidence for the prevention and treatment of DD in this population. Data from the Global Burden of Disease (GBD) 2021 were used to study DD in women aged 45–54 years. The samples were split into two groups (45–49 and 50–54) for analysis. Three metrics—incidence, prevalence, and disability-adjusted life year (DALY)—were assessed to evaluate the disease burden of DD in menopausal women. Between 1990 and 2021, the age-standardized incidence rate (ASIR), age-standardized prevalence rate (ASPR), and age-standardized DALY rate (ASDR) of DD in menopausal women aged 45–49 years globally declined, with estimated annual percentage changes (EAPCs) of -0.14 [-0.25, -0.02], -0.17 [-0.34, -0.01], and -0.16 [-0.29, -0.02], respectively. Conversely, no significant changes were detected in women aged 50–54 years (EAPCs: -0.08 [-0.18, 0.02], -0.13 [-0.26, -0.00], and -0.10 [-0.22, 0.01]). Regionally, in high-SDI areas, the ASPR, ASIR, and ASDR of DD in women aged 45–49 years increased (EAPCs: 0.30 [0.19, 0.42], 0.50 [0.32, 0.69], 0.39 [0.24, 0.54]), whereas those in low-, low-, middle-, middle-, and high- to middle-SDI regions decreased. Similarly, in high-SDI regions, the ASPR, ASIR, and ASDR of DD in women aged 50–54 rose (EAPCs: 0.36 [0.26, 0.45], 0.49 [0.34, 0.65], 0.41 [0.29, 0.54]), whereas other SDI regions showed a decline. This study analyzed the burden of DD among menopausal women aged 45--54 years globally from 1990--2021, revealing trend changes at the global, national, and regional levels. Despite a significant increase in the burden of DD among menopausal women over the past 31 years, the rates of increase in ASPR, ASIR, and ASDR have not continued to rise. Significant trend differences were observed across regions and age groups. Additionally, high-SDI regions experienced a faster increase in the burden of DD, whereas low-SDI regions faced a greater disease burden.

## Introduction

Depressive disorders (DD) are the most prevalent and disabling mental health conditions, affecting approximately 15–18% of the general population globally [1–3]. Most women experience menopause between the ages of 45 and 55[4], a natural part of physiological aging. Hormonal changes associated with menopause can impact physical, emotional, psychological, and social well-being, including symptoms such as mood swings, depression, and anxiety [5]. The global prevalence of DD in women is increasing, particularly during the menopausal period[6–8]. DD in menopausal women not only affects their quality of life but can also lead to long-term health issues, such as chronic disease and cognitive decline[9]. Some studies have shown that the incidence of DD in postmenopausal women is significantly greater than that in perimenopausal women[10, 11], with women in the 50--54 age group being mostly postmenopausal[4, 12]. Postmenopausal women often experience more concentrated menopausal symptoms, including hot flashes, mood swings, and sexual dysfunction[13]. The risk of developing cardiovascular diseases and osteoporosis is also significantly increased[9]. Moreover, these symptoms tend to be more persistent in postmenopausal women [14].

Understanding the epidemiological trends of DD in menopausal women is crucial for improving their quality of life during this period. However, studies on the global burden of DD in menopausal women across different regions and age groups are limited. This study leverages data from the Global Burden of Disease (GBD) 2021 to analyze the incidence rate, prevalence rate, and disability-adjusted life years (DALY) of DD among women aged 45--54 years from 1990--2021. These findings enhance our understanding of the global burden of DD in menopausal women and provide guidance for policy-making.

## Methods

### Data Source

Data were extracted from the GBD database website (http://ghdx.healthdata.org), which contains data on various diseases from 204 countries and regions. Data on the incidence rate, prevalence rate, and DALYs of DD among women aged 45--54 years from 1990--2021 were analyzed to comprehensively describe the disease burden of DD.

### Global and Regional Burden Analysis

To assess the global distribution and regional differences in the disease burden of DD among menopausal women, global and regional maps were created via R software (version 4.3.3) and the JD GBDR package (V2.22, Jingding Medical Technology Co., Ltd.).

### Trend Analysis

Join-point regression analysis was employed to evaluate the time trends of the incidence rate, prevalence rate, mortality rate, and DALYs of DD from 1990--2021. The analysis was conducted via the Segment and broom R packages, which identify significant trend changes over time. The estimated annual percentage changes (EAPCs) and their 95% confidence intervals (CIs) were used to determine the statistical significance of the trends.

### Sociodemographic Index (SDI) Analysis

The GBD database categorizes 204 countries and regions into five SDI levels: high-middle-SDI (≥0.81), high-middle-SDI (0.70--0.81), middle-SDI (0.61--0.69), low-middle-SDI (0.46--0.60), and low-SDI (<0.46). Data were analyzed to compare the disease burden across regions with different levels of sociodemographic development via the dplyr and ggplot2 packages in R.

### Statistical analysis

All the statistical analyses and data visualizations were performed via R (version 4.3.3) and JD GBDR (V2.24; Jingding Medical Technology Co., Ltd.). The age-standardized rate (ASR) and EAPCs were used to assess the incidence trends of DD. ASRs were adjusted for age-related variations to eliminate the confounding effects of age structure on incidence rates. Global maps and regional comparisons were generated to analyze the global distribution and regional differences.

### Ethical Considerations

The data for this study were drawn from the Global Burden of Disease (GBD) 2021 database. This database is publicly available and does not involve direct contact with human participants. Given this, formal ethical approval and individual informed consent were not necessary. However, we are fully aware of the ethical responsibilities that come with using human health data. We have made sure to use the data in line with the GBD study’s guidelines and regulations. Our study also follows the principles of the Declaration of Helsinki, which sets the standard for ethical research conduct.

## Results

### Global trends

Globally, the estimated prevalence of depressive disorder (DD) among menopausal women aged 45–49 years increased from 79.32×10^5^ cases in 1990 (95% CI: 67.39×10^5^–93.75×10^5^) to 177.43×10^5^ cases in 2021 (95% CI: 149.56×10^5^–210.24×10^5^), representing a 123.69% increase over the 31-year period (Table 1, Supplementary Figure 1). Similarly, for women aged 50--54 years, the estimated prevalence of DD rose from 73.7×10^5^ cases in 1990 (95% CI: 64.02×10^5^– 84.79×10^5^) to 169.56×10^5^ cases in 2021 (95% CI: 146.08×10^5^–196.60×10^5^), an increase of 130.07% (Table 2, Supplementary Figure 1). However, a notable discrepancy was observed between the estimated case numbers and the age-standardized prevalence rates (ASPRs) over the three decades. Specifically, the ASPR for the 45–49 age group exhibited a declining trend (EAPCs = -0.14, 95% CI: -0.25, -0.02), despite the significant increase in case numbers (Table 1). Similarly, the ASPR for the 50–54 age group did not significantly change (EAPCs = -0.08, 95% CI: -0.18, 0.02) despite the increase in prevalence (Table 2).

**Table 1.** Prevalence incidence and disability-adjusted life years of depression in menopausal women aged 45-49 years, from 1990 to 2021.

|  |  |  |  |  |  |  |
| --- | --- | --- | --- | --- | --- | --- |
| Type | 1990 |  | 2021 |  | EAPCs (95% CI) | Percent change in number (%) |
|  | Number No×10<sup>5</sup> (95% CI) | ASPR per 100000(95% CI) | Number No×10<sup>5</sup> (95% CI) | ASPR per 100000(95% CI) |  |  |
| Prevalence |  |  |  |  |  |  |
| Global | 79.32 ( 93.75 , 67.39 ) | 6970.24 ( 8238.23 , 5921.91 ) | 177.43 ( 210.24 , 149.56 ) | 7529.49 ( 8921.84 , 6347.01 ) | -0.14(-0.25, -0.02) | 123.68(118.47,128.83) |
| SDI Region |  |  |  |  |  |  |
| Low SDI | 7.13 ( 8.6, 5.81 ) | 8594.49 ( 10358.88, 7003 ) | 18.26 ( 22.06, 14.87 ) | 8794.94 ( 10621.61, 7161.91 ) | -0.28(-0.40, -0.15) | 155.99(147.04,166.01) |
| Low-middle SDI | 18.07 ( 21.41, 15.01 ) | 8327.03 ( 9863.24, 6913.97 ) | 42.76 ( 51.47, 35.35 ) | 8650.11 ( 10411.44, 7149.66 ) | -0.44(-0.62, -0.26) | 136.60(126.69,148.15) |
| Middle SDI | 22.64 ( 26.82, 19.26 ) | 6679.54 ( 7913.5, 5682.35 ) | 56.93 ( 66.99, 47.97 ) | 7018.66 ( 8258.99, 5914.2 ) | -0.22(-0.33, -0.11) | 151.46(143.86,159.27) |
| High-middle SDI | 16.57 ( 19.48, 14.14 ) | 6759.12 ( 7946, 5767.89 ) | 33.83 ( 40.05, 28.46 ) | 7014.42 ( 8304.7, 5901.36 ) | -0.21(-0.31, -0.11) | 104.14(95.60,111.90) |
| High SDI | 14.83 ( 17.29, 12.75 ) | 5868.32 ( 6844.24, 5044.41 ) | 25.51 ( 29.86, 21.61 ) | 7103.29 ( 8315.25, 6019.24 ) | 0.30(0.19,0.42) | 72.01(64.12,81.51) |
| GBD Region |  |  |  |  |  |  |
| Andean Latin America | 0.38 ( 0.47 , 0.3 ) | 5347.71 ( 6657 , 4239.39 ) | 1.16 ( 1.44 , 0.91 ) | 6197.85 ( 7692.39 , 4876.52 ) | 0.04(-0.16,0.24) | 205.01(174.17,243.14) |
| Australasia | 0.38 ( 0.46 , 0.32 ) | 6745.1 ( 8148.49 , 5574.42 ) | 0.69 ( 0.88 , 0.53 ) | 6842.52 ( 8770.27 , 5295.21 ) | -0.10(-0.30,0.10) | 80.58(47.72,121.16) |
| Caribbean | 0.61 ( 0.73 , 0.49 ) | 7664.1 ( 9279.3 , 6190.78 ) | 1.13 ( 1.41 , 0.88 ) | 7795.47 ( 9744.15 , 6082.17 ) | -0.32(-0.47, -0.17) | 86.99(67.67,108.35) |
| Central Asia | 0.74 ( 0.9 , 0.6 ) | 6307.17 ( 7709.62 , 5159.71 ) | 1.86 ( 2.3 , 1.49 ) | 6766.03 ( 8357.98 , 5424.8 ) | -0.02(-0.13,0.08) | 151.82(132.95,175.57) |
| Central Europe | 2 ( 2.41 , 1.65 ) | 5858.7 ( 7071.64 , 4849.51 ) | 2.64 ( 3.21 , 2.18 ) | 6163.76 ( 7486.7 , 5083.6 ) | -0.25(-0.41, -0.09) | 32.22(24.80,40.58) |
| Central Latin America | 1.73 ( 2.11 , 1.39 ) | 5783.86 ( 7038.25 , 4659.2 ) | 5.9 ( 7.2 , 4.72 ) | 7441.11 ( 9078.12 , 5951.83 ) | 0.58(0.45,0.71) | 240.79(224.05,258.18) |
| Central Sub-Saharan Africa | 0.93 ( 1.16 , 0.73 ) | 10347.34 ( 12876.98 , 8158.35 ) | 2.69 ( 3.46 , 2.08 ) | 10822.61 ( 13926.82 , 8362.26 ) | 0.05(-0.02,0.11) | 189.13(152.80,231.23) |
| East Asia | 16.48 ( 19.58 , 13.95 ) | 6470.77 ( 7691.71 , 5479.49 ) | 34.8 ( 41.52 , 29.03 ) | 6199.39 ( 7397.6 , 5171.19 ) | -0.39(-0.46, -0.32) | 111.21(96.05,123.87) |
| Eastern Europe | 3.94 ( 4.7 , 3.25 ) | 6980.94 ( 8333.71 , 5756.35 ) | 5.63 ( 6.78 , 4.58 ) | 7436.48 ( 8955.33 , 6047.8 ) | -0.31(-0.46, -0.17) | 42.91(36.53,50.87) |
| Eastern Sub-Saharan Africa | 2.73 ( 3.28 , 2.23 ) | 9467.96 ( 11392.81 , 7754.93 ) | 7.31 ( 8.89 , 5.97 ) | 9706.72 ( 11794.63 , 7923.17 ) | -0.19(-0.28, -0.11) | 168.20(155.33,182.82) |
| High-income Asia Pacific | 1.88 ( 2.21 , 1.6 ) | 3241.58 ( 3805.9 , 2765.1 ) | 2.63 ( 3.14 , 2.22 ) | 9401.98 ( 11744.06 , 7254.08 ) | 0.19(0.03,0.34) | 39.60(31.49,48.68) |
| High-income North America | 5.35 ( 6.27 , 4.57 ) | 6752.01 ( 7921.89 , 5772.16 ) | 10.15 ( 11.94 , 8.66 ) | 5700.82 ( 7417.64 , 4380.56 ) | 0.44(0.28,0.60) | 89.72(75.87,105.69) |
| North Africa and Middle East | 4.93 ( 6.01 , 3.96 ) | 8525.64 ( 10404.31 , 6854.8 ) | 15.88 ( 19.83 , 12.25 ) | 8681.2 ( 10272.83 , 7249.24 ) | 0.18(0.10,0.26) | 222.28(202.46,241.63) |
| Oceania | 0.06 ( 0.08 , 0.05 ) | 5589.7 ( 7299.62 , 4333.19 ) | 0.18 ( 0.24 , 0.14 ) | 5634.14 ( 6947.13 , 4650.11 ) | -0.02(-0.04,0.01) | 187.10(162.54,215.95) |
| South Asia | 17.72 ( 20.75 , 14.78 ) | 8547.55 ( 10009.01 , 7131.66 ) | 42.77 ( 50.61 , 35.72 ) | 5678.16 ( 7061.25 , 4554.9 ) | -0.68(-0.92, -0.43) | 141.42(129.80,154.60) |
| Southeast Asia | 4.91 ( 6.11 , 4.01 ) | 5331.62 ( 6628.33 , 4353.5 ) | 12.56 ( 15.48 , 10.36 ) | 9455.59 ( 11206.5 , 7936.61 ) | -0.03(-0.10,0.03) | 155.67(148.45,164.13) |
| Southern Latin America | 0.69 ( 0.83 , 0.57 ) | 5345.21 ( 6470.27 , 4417.97 ) | 1.24 ( 1.54 , 1 ) | 9048.41 ( 10616.91 , 7519.21 ) | -0.22(-0.37, -0.08) | 80.87(57.47,108.01) |
| Southern Sub-Saharan Africa | 0.79 ( 0.94 , 0.67 ) | 8349.06 ( 9918.59 , 7042.04 ) | 1.99 ( 2.35 , 1.67 ) | 8357.58 ( 10031.12 , 6886.84 ) | 0.14(-0.02,0.30) | 150.06(135.21,167.50) |
| Tropical Latin America | 2.65 ( 3.1 , 2.24 ) | 8295.33 ( 9720.87 , 7011.28 ) | 6.81 ( 7.99 , 5.66 ) | 7906.63 ( 9537.33 , 6428.01 ) | -0.30(-0.56, -0.05) | 157.43(140.60,174.97) |
| Western Europe | 8.03 ( 9.38 , 6.9 ) | 7079.6 ( 8271.95 , 6084.67 ) | 12.47 ( 14.96 , 10.27 ) | 9401.98 ( 11744.06 , 7254.08 ) | 0.26(0.14,0.38) | 55.30(43.30,69.01) |
| Western Sub-Saharan Africa | 2.42 ( 2.92 , 1.99 ) | 8158.98 ( 9860.85 , 6721.4 ) | 6.95 ( 8.38 , 5.65 ) | 5700.82 ( 7417.64 , 4380.56 ) | -0.24(-0.31,-0.17) | 187.72(178.46,197.26) |
| incidence |  |  |  |  |  |  |
| Global | 78.18 ( 94.1 , 63.67 ) | 6870.15 ( 8268.45 , 5594.54 ) | 180.6 ( 216.86 , 146.68 ) | 7663.94 ( 9203.11 , 6224.61 ) | -0.17(-0.34,-0.01) | 130.99(125.04,136.70) |
| **SDI Region** |  |  |  |  |  |  |
| Low SDI | 7.75 ( 9.65 , 6.1 ) | 9334.13 ( 11623.54 , 7351.11 ) | 20.03 ( 24.51 , 15.85 ) | 9647.06 ( 11803.46 , 7630.85 ) | -0.38(-0.55,-0.21) | 158.55(145.75,172.75) |
| Low-middle SDI | 19.75 ( 24.16 , 15.85 ) | 9101.17 ( 11129.09 , 7304.57 ) | 47.4 ( 57.76 , 37.79 ) | 9587.83 ( 11683.67 , 7643.61 ) | -0.60(-0.84,-0.36) | 139.95(127.07,154.62) |
| Middle SDI | 20.42 ( 24.67 , 16.64 ) | 6023.74 ( 7279.46 , 4909.36 ) | 54.07 ( 65.22 , 43.92 ) | 6666.19 ( 8041.23 , 5414.39 ) | -0.18(-0.35,-0.01) | 164.84(156.27,173.54) |
| High-middle SDI | 15.6 ( 18.69 , 12.8 ) | 6362.41 ( 7624.19 , 5222.1 ) | 31.61 ( 38.72 , 25.35 ) | 6553.26 ( 8027.85 , 5255.93 ) | -0.32(-0.48,-0.16) | 102.61(91.59,112.83) |
| High SDI | 14.59 ( 17.38 , 12.25 ) | 5774.04 ( 6876.86 , 4850.12 ) | 27.35 ( 33.44 , 22.59 ) | 7615.94 ( 9312.69 , 6291.02 ) | 0.50(0.32,0.69) | 87.44(77.05,97.68) |
| **GBD Region** |  |  |  |  |  |  |
| Andean Latin America | 0.4 ( 0.51 , 0.3 ) | 5601.09 ( 7132.09 , 4283.89 ) | 1.29 ( 1.7 , 0.97 ) | 6909.72 ( 9070.43 , 5175.19 ) | 0.06(-0.22,0.34) | 224.66(180.16,277.75) |
| Australasia | 0.41 ( 0.51 , 0.34 ) | 7327.67 ( 8997 , 6036.71 ) | 0.75 ( 1.02 , 0.55 ) | 7454.9 ( 10114.41 , 5497.43 ) | -0.13(-0.38,0.12) | 81.10(39.16,133.29) |
| Caribbean | 0.71 ( 0.88 , 0.56 ) | 9001.73 ( 11186.03 , 7095.22 ) | 1.35 ( 1.72 , 1.01 ) | 9279.95 ( 11852.11 , 6979.19 ) | -0.38(-0.57,-0.19) | 89.52(64.50,117.02) |
| Central Asia | 0.76 ( 0.93 , 0.61 ) | 6455.41 ( 7982.48 , 5183.55 ) | 1.96 ( 2.51 , 1.48 ) | 7153.9 ( 9156.29 , 5383.14 ) | -0.05(-0.20,0.11) | 160.14(132.11,197.71) |
| Central Europe | 1.94 ( 2.36 , 1.58 ) | 5677.56 ( 6909.17 , 4639.77 ) | 2.66 ( 3.24 , 2.11 ) | 6201.16 ( 7566.76 , 4922.14 ) | -0.37(-0.63,-0.11) | 37.27(25.29,49.63) |
| Central Latin America | 1.93 ( 2.43 , 1.51 ) | 6440.42 ( 8100.43 , 5034.03 ) | 7.01 ( 8.81 , 5.47 ) | 8839.23 ( 11106.36 , 6901.77 ) | 0.72(0.56,0.88) | 263.55(242.69,285.57) |
| Central Sub-Saharan Africa | 1.05 ( 1.37 , 0.8 ) | 11675.26 ( 15197.39 , 8910.12 ) | 3.08 ( 4.09 , 2.27 ) | 12402.31 ( 16457.15 , 9133.04 ) | 0.06(-0.02,0.15) | 193.65(147.80,251.00) |
| East Asia | 12.18 ( 14.77 , 10.02 ) | 4785.59 ( 5799.73 , 3936.92 ) | 25.14 ( 30.27 , 20.49 ) | 4478.16 ( 5393 , 3649.94 ) | -0.34(-0.54,-0.14) | 106.29(90.01,121.49) |
| Eastern Europe | 4.13 ( 5.05 , 3.24 ) | 7312.16 ( 8948.28 , 5750.45 ) | 6.05 ( 7.63 , 4.68 ) | 7987.65 ( 10077.31 , 6187.13 ) | -0.47(-0.69,-0.26) | 46.55(37.47,58.57) |
| Eastern Sub-Saharan Africa | 2.98 ( 3.71 , 2.37 ) | 10353.43 ( 12885.86 , 8211.49 ) | 8.15 ( 10.1 , 6.4 ) | 10821.26 ( 13404.72 , 8493.49 ) | -0.24(-0.36,-0.11) | 173.43(156.31,194.61) |
| High-income Asia Pacific | 1.9 ( 2.29 , 1.56 ) | 3277.98 ( 3952.29 , 2692.59 ) | 2.79 ( 3.42 , 2.25 ) | 3885.49 ( 4763.51 , 3138.6 ) | 0.41(0.23,0.59) | 46.55(33.97,59.34) |
| High-income North America | 4.72 ( 5.72 , 3.93 ) | 5958.08 ( 7217.22 , 4962.8 ) | 11.07 ( 13.41 , 9.2 ) | 9909.32 ( 12001.22 , 8236.75 ) | 0.91(0.58,1.24) | 134.58(119.31,151.39) |
| North Africa and Middle East | 5.44 ( 6.81 , 4.21 ) | 9417.56 ( 11779.48 , 7294.53 ) | 18.12 ( 23.31 , 13.65 ) | 10733.02 ( 13805.62 , 8081.78 ) | 0.23(0.13,0.34) | 233.07(209.96,258.26) |
| Oceania | 0.05 ( 0.06 , 0.03 ) | 3952.5 ( 5015.45 , 3041.96 ) | 0.13 ( 0.18 , 0.1 ) | 4126.1 ( 5630.81 , 3011.41 ) | -0.03(-0.09,0.02) | 193.87(144.24,255.28) |
| South Asia | 19.98 ( 24.21 , 16.17 ) | 9638.77 ( 11682.7 , 7800.66 ) | 48.36 ( 58.28 , 39.05 ) | 9815.91 ( 11828.02 , 7925.64 ) | -0.91(-1.23,-0.59) | 142.07(128.74,156.47) |
| Southeast Asia | 3.29 ( 4.02 , 2.64 ) | 3570.54 ( 4367.55 , 2862.23 ) | 9.01 ( 11.23 , 6.98 ) | 4041.28 ( 5036.37 , 3133.2 ) | -0.08(-0.22,0.07) | 173.83(158.14,189.80) |
| Southern Latin America | 0.78 ( 0.98 , 0.62 ) | 6077.04 ( 7622.23 , 4865.39 ) | 1.44 ( 1.86 , 1.11 ) | 6577.42 ( 8532.47 , 5072.49 ) | -0.16(-0.32,0.00) | 84.29(54.04,118.45) |
| Southern Sub-Saharan Africa | 0.83 ( 1.01 , 0.67 ) | 8717.08 ( 10614.24 , 7091.46 ) | 2.18 ( 2.67 , 1.75 ) | 10375.76 ( 12682.55 , 8307.27 ) | 0.18(-0.06,0.41) | 162.82(141.55,186.41) |
| Tropical Latin America | 3.18 ( 3.82 , 2.61 ) | 9976.5 ( 11987.46 , 8191.45 ) | 8.31 ( 10.1 , 6.71 ) | 11042.9 ( 13421.78 , 8918.89 ) | -0.38(-0.70,-0.06) | 161.24(140.63,181.96) |
| Western Europe | 9.01 ( 10.72 , 7.54 ) | 7947.67 ( 9450.86 , 6648.1 ) | 14.59 ( 17.94 , 11.69 ) | 9782.11 ( 12025.25 , 7837.31 ) | 0.31(0.15,0.46) | 61.92(46.80,79.52) |
| Western Sub-Saharan Africa | 2.52 ( 3.11 , 2 ) | 8510.03 ( 10493.39 , 6761.76 ) | 7.15 ( 8.85 , 5.68 ) | 8133.27 ( 10068.95 , 6459.09 ) | -0.35(-0.45,-0.24) | 183.75(169.81,197.80) |
| **DALY** |  |  |  |  |  |  |
| Global | 12.97 ( 17.87 , 8.66 ) | 1139.93 ( 1570.52 , 760.58 ) | 29.38 ( 40.63 , 19.4 ) | 1246.69 ( 1724.17 , 823.39 ) | -0.16(-0.29,-0.02) | 126.46(120.55,131.48) |
| **SDI Region** |  |  |  |  |  |  |
| Low SDI | 1.21 ( 1.68 , 0.79 ) | 1451.79 ( 2028.44 , 948.64 ) | 3.11 ( 4.34 , 2.04 ) | 1496.79 ( 2089.48 , 980.86 ) | -0.31(-0.45,-0.16) | 157.91(145.80,170.06) |
| Low-middle SDI | 3.07 ( 4.26 , 2 ) | 1416.34 ( 1962.08 , 920.91 ) | 7.32 ( 10.25 , 4.75 ) | 1481.48 ( 2073.16 , 959.97 ) | -0.51(-0.72,-0.30) | 138.24(126.29,152.46) |
| Middle SDI | 3.56 ( 4.93 , 2.38 ) | 1051.77 ( 1454.79 , 701.83 ) | 9.13 ( 12.62 , 6.06 ) | 1125.72 ( 1556.05 , 747.02 ) | -0.21(-0.34,-0.08) | 156.14(147.18,164.15) |
| High-middle SDI | 2.67 ( 3.67 , 1.79 ) | 1088.41 ( 1497.61 , 729.24 ) | 5.42 ( 7.57 , 3.62 ) | 1124.28 ( 1569.22 , 750.02 ) | -0.26(-0.38,-0.13) | 103.19(92.39,112.81) |
| High SDI | 2.45 ( 3.35 , 1.68 ) | 968.65 ( 1327.34 , 664.11 ) | 4.37 ( 6.08 , 2.97 ) | 1216.87 ( 1691.92 , 828.4 ) | 0.39(0.24,0.54) | 78.53(69.21,88.71) |
| **GBD Region** |  |  |  |  |  |  |
| Andean Latin America | 0.06 ( 0.09 , 0.04 ) | 898.04 ( 1315.63 , 556.99 ) | 0.2 ( 0.29 , 0.13 ) | 1072.09 ( 1575.35 , 678.63 ) | 0.05(-0.19,0.29) | 214.18(171.85,263.63) |
| Australasia | 0.07 ( 0.09 , 0.04 ) | 1166.55 ( 1647.14 , 760.04 ) | 0.12 ( 0.18 , 0.07 ) | 1190.28 ( 1762.37 , 733.05 ) | -0.11(-0.34,0.12) | 81.63(40.01,135.37) |
| Caribbean | 0.11 ( 0.16 , 0.07 ) | 1366.35 ( 1966.6 , 889.63 ) | 0.2 ( 0.29 , 0.13 ) | 1390.58 ( 2028.4 , 874.13 ) | -0.37(-0.54,-0.20) | 87.10(62.42,110.79) |
| Central Asia | 0.12 ( 0.17 , 0.08 ) | 1040.85 ( 1461.13 , 677.48 ) | 0.31 ( 0.44 , 0.2 ) | 1134.23 ( 1613.5 , 722.57 ) | -0.02(-0.15,0.10) | 155.80(127.62,184.58) |
| Central Europe | 0.32 ( 0.45 , 0.21 ) | 946.13 ( 1317.38 , 617.05 ) | 0.43 ( 0.61 , 0.28 ) | 1010.56 ( 1413.79 , 665.11 ) | -0.31(-0.51,-0.10) | 34.24(24.05,46.32) |
| Central Latin America | 0.3 ( 0.43 , 0.19 ) | 998.95 ( 1434.62 , 637.17 ) | 1.05 ( 1.52 , 0.66 ) | 1329.81 ( 1918.61 , 832.27 ) | 0.66(0.52,0.80) | 252.62(231.86,275.39) |
| Central Sub-Saharan Africa | 0.16 ( 0.23 , 0.1 ) | 1786.83 ( 2564.48 , 1131.14 ) | 0.47 ( 0.7 , 0.29 ) | 1897.94 ( 2806.27 , 1150.52 ) | 0.09(0.02,0.17) | 193.63(150.62,245.58) |
| East Asia | 2.42 ( 3.34 , 1.65 ) | 950.32 ( 1313.38 , 646.68 ) | 5.03 ( 6.87 , 3.48 ) | 896.1 ( 1224.14 , 619.76 ) | -0.39(-0.51,-0.28) | 107.87(93.30,121.18) |
| Eastern Europe | 0.66 ( 0.92 , 0.43 ) | 1165.77 ( 1631.03 , 762.87 ) | 0.95 ( 1.36 , 0.63 ) | 1256.84 ( 1799.99 , 833.81 ) | -0.38(-0.56,-0.20) | 44.64(35.52,55.65) |
| Eastern Sub-Saharan Africa | 0.46 ( 0.65 , 0.3 ) | 1609.54 ( 2243.09 , 1053.67 ) | 1.25 ( 1.78 , 0.81 ) | 1665.03 ( 2366.99 , 1070.58 ) | -0.20(-0.30,-0.10) | 170.63(155.55,189.42) |
| High-income Asia Pacific | 0.32 ( 0.44 , 0.21 ) | 543.9 ( 756.91 , 362.77 ) | 0.45 ( 0.63 , 0.3 ) | 630.42 ( 874.04 , 415.62 ) | 0.30(0.14,0.47) | 43.31(31.03,55.37) |
| High-income North America | 0.84 ( 1.16 , 0.58 ) | 1064.47 ( 1460.19 , 731.53 ) | 1.75 ( 2.44 , 1.2 ) | 1565.17 ( 2186.48 , 1072.35 ) | 0.63(0.40,0.86) | 107.38(91.32,124.46) |
| North Africa and Middle East | 0.85 ( 1.22 , 0.55 ) | 1471.35 ( 2109.13 , 948.57 ) | 2.77 ( 4.01 , 1.74 ) | 1639.32 ( 2377.44 , 1029.21 ) | 0.20(0.11,0.29) | 225.61(205.40,246.77) |
| Oceania | 0.01 ( 0.01 , 0.01 ) | 792.98 ( 1147.33 , 501.7 ) | 0.03 ( 0.04 , 0.02 ) | 817.26 ( 1215.99 , 508.12 ) | -0.02(-0.05,0.02) | 190.13(147.82,234.61) |
| South Asia | 3.05 ( 4.23 , 2 ) | 1469.73 ( 2042.86 , 966.97 ) | 7.37 ( 10.3 , 4.82 ) | 1496.12 ( 2091.43 , 977.85 ) | -0.79(-1.07,-0.50) | 141.98(128.00,156.66) |
| Southeast Asia | 0.69 ( 0.97 , 0.45 ) | 746.67 ( 1058.28 , 493.44 ) | 1.81 ( 2.55 , 1.2 ) | 811.18 ( 1146.05 , 540.16 ) | -0.04(-0.13,0.05) | 162.84(150.57,177.02) |
| Southern Latin America | 0.12 ( 0.17 , 0.08 ) | 937.96 ( 1314.49 , 619.56 ) | 0.22 ( 0.32 , 0.14 ) | 1006.25 ( 1447.7 , 642.04 ) | -0.18(-0.34,-0.03) | 82.66(51.51,117.97) |
| Southern Sub-Saharan Africa | 0.13 ( 0.18 , 0.09 ) | 1395.47 ( 1936.71 , 907.09 ) | 0.33 ( 0.46 , 0.22 ) | 1584.33 ( 2207.74 , 1042.73 ) | 0.10(-0.10,0.29) | 150.68(132.30,171.94) |
| Tropical Latin America | 0.47 ( 0.66 , 0.32 ) | 1479.52 ( 2076.79 , 990.73 ) | 1.23 ( 1.72 , 0.8 ) | 1630.11 ( 2287.58 , 1065.27 ) | -0.33(-0.62,-0.05) | 160.03(140.81,180.78) |
| Western Europe | 1.41 ( 1.95 , 0.96 ) | 1244.82 ( 1721.39 , 846.21 ) | 2.24 ( 3.17 , 1.51 ) | 1503.15 ( 2122.37 , 1013.13 ) | 0.29(0.15,0.43) | 58.85(44.63,73.71) |
| Western Sub-Saharan Africa | 0.4 ( 0.56 , 0.26 ) | 1354.79 ( 1900.12 , 883.67 ) | 1.15 ( 1.63 , 0.74 ) | 1310.41 ( 1851.59 , 845.81 ) | -0.27(-0.35,-0.18) | 187.17(175.17,200.21) |

**Table 2.**
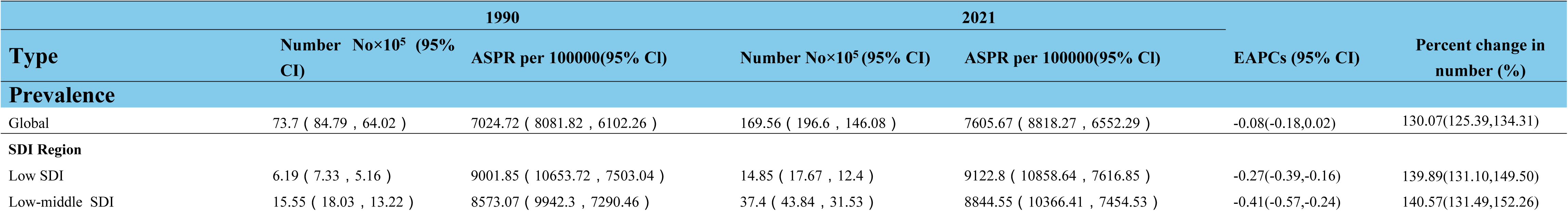
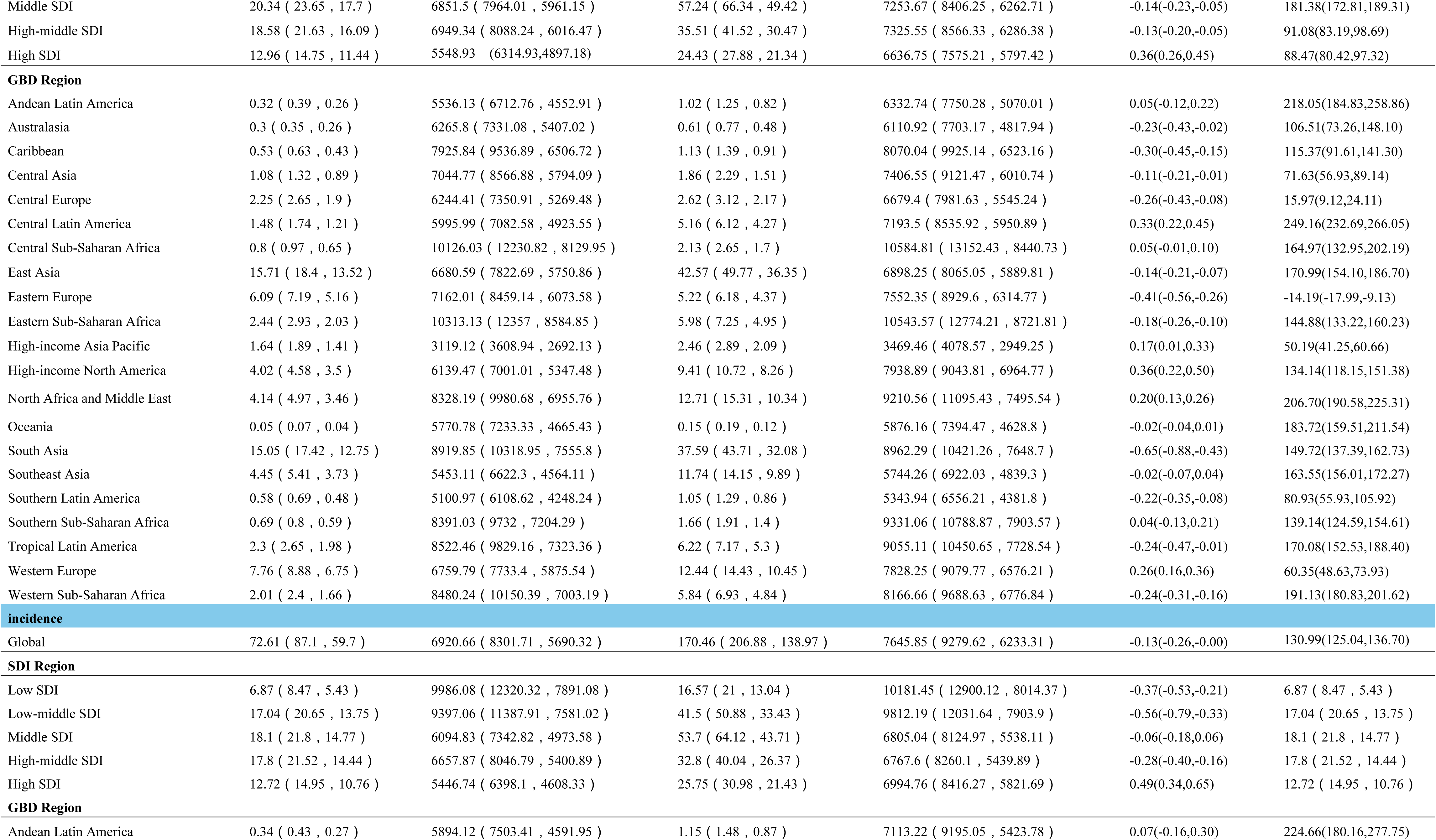
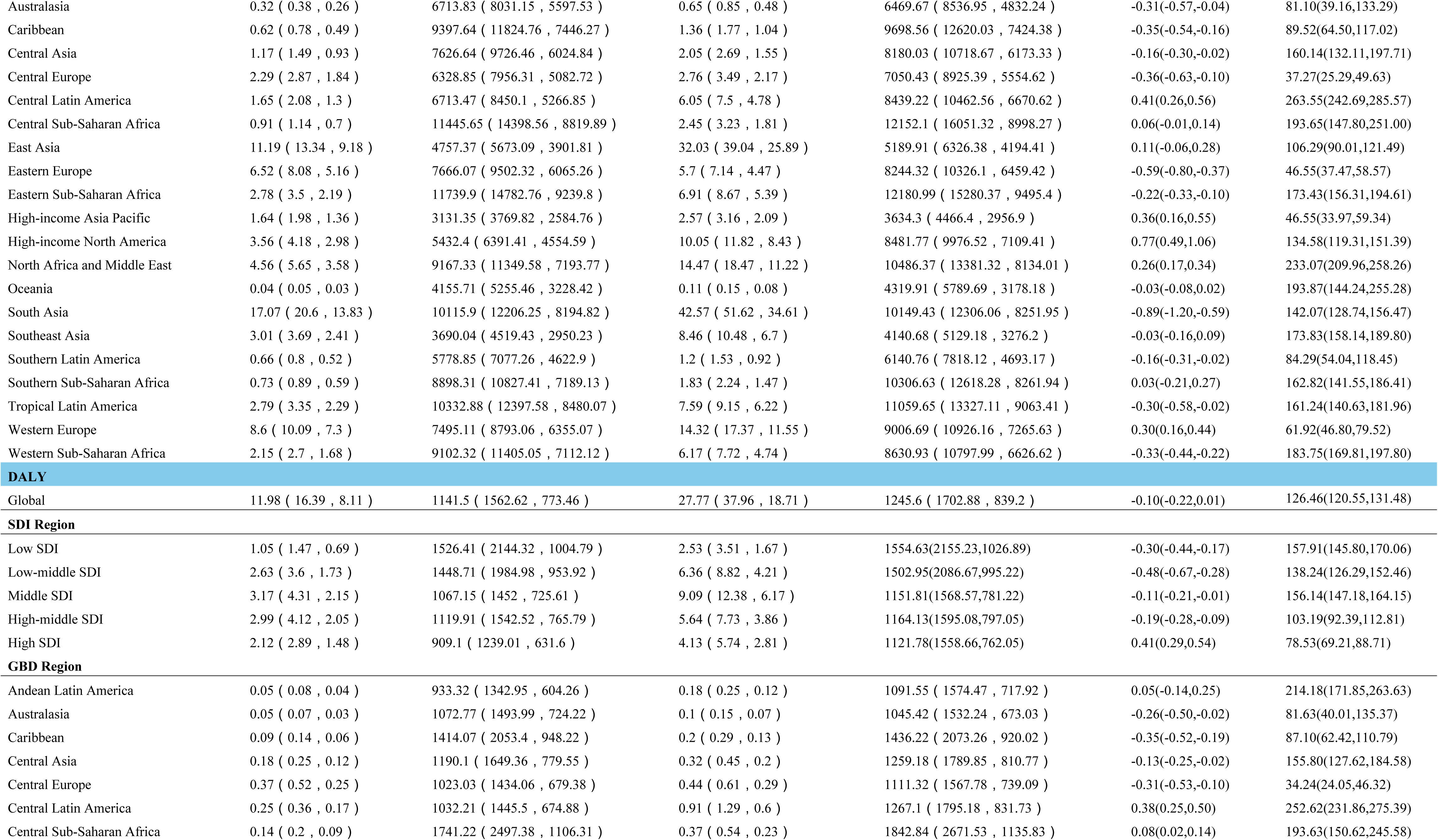
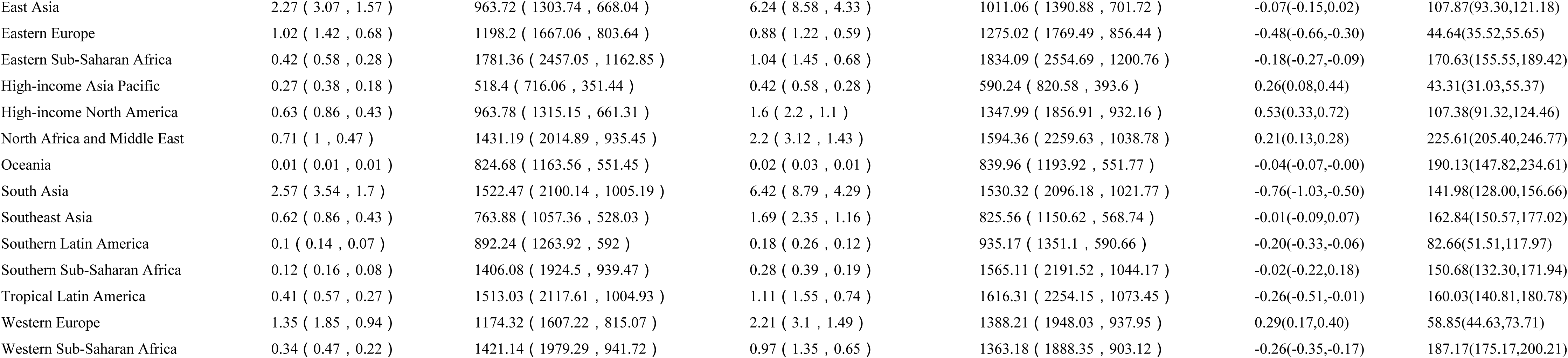
Prevalence incidence and disability-adjusted life years of depression in menopausal women aged 50-54 years, from 1990 to 2021.

This inconsistency was also evident in the trends of incidence rates. The estimated incidence of DD in the 45–49 years age group increased from 78.18×10^5^ cases in 1990 (95% CI: 63.67×10^5^–94.10×10^5^) to 180.60×10^5^ cases in 2021 (95% CI: 146.68×10^5^–216.86×10^5^), a 130.99% increase (95% CI: 125.04%, 136.70%) (Supplementary Figure 2). For the 50–54 year age group, the estimated incidence of DD increased from 72.61×10^5^ cases in 1990 (95% CI: 59.70×10^5^– 87.10×10^5^) to 170.46×10^5^ cases in 2021 (95% CI: 138.97×10^5^–206.88×10^5^), which was also a 130.99% increase (95% CI: 125.04%, 136.70%) (Supplementary Figure 2). Despite these increases, the age-standardized incidence rate (ASIR) for the 45–49 years age group showed a declining trend (EAPCs = -0.17, 95% CI: -0.34, -0.01), whereas the trend for the 50–54 years age group was not significant (EAPCs = -0.13, 95% CI: -0.26, -0.00). (Table 1. Table 2).

The disability-adjusted life years (DALYs) associated with DD also increased in both age groups. For the 45–49 age group, DALYs rose from 12.97×10^5^ in 1990 (95% CI: 8.66×10^5^–17.87×10^5^) to 29.38×10^5^ in 2021 (95% CI: 19.40×10^5^–40.63×10^5^), a 126.46% increase (95% CI: 120.55%, 131.48%) (Supplementary Figure 3). For the 50–54 age group, DALYs increased from 11.98×10^5^ in 1990 (95% CI: 8.11×10^5^–16.39×10^5^) to 27.77×10^5^ in 2021 (95% CI: 18.71×10^5^–37.96×10^5^), which was also a 126.46% increase (95% CI: 120.55%, 131.48%) (Supplementary Figure 3). However, the age-standardized DALY rate (ASDR) for the 45–49 years age group showed a declining trend (EAPCs = -0.16, 95% CI: -0.29, -0.02), whereas the trend for the 50–54 years age group was not significant (EAPCs = -0.10, 95% CI: -0.22, 0.01).(Table 1. Table 2).

### SDI level analysis

In 1990, the ASPR, ASIR, and ASDR for menopausal women aged 45–49 years were highest in low-SDI regions, with values of 8594.49 per 100,000 (95% CI: 7003–10358.88), 9334.13 per 100,000 (95% CI: 7351.11–11623.54), and 1451.79 per 100,000 (95% CI: 948.64–2028.44), respectively (Table 1). Similarly, for women aged 50–54 years, the ASPR, ASIR, and ASDR were also highest in low-SDI regions, with values of 9001.85 per 100,000 (95% CI: 7503.04–10653.72), 9986.08 per 100,000 (95% CI: 7891.08–12320.32), and 1526.41 per 100,000 (95% CI: 1004.79–2144.32), respectively (Table 2).

By 2021, the ASPR, ASIR, and ASDR for menopausal women aged 45–49 years remained highest in low-SDI regions, with values of 8794.94 per 100,000 (95% CI: 7161.91–10621.61), 9647.06 per 100,000 (95% CI: 7630.85–11803.46), and 1496.79 per 100,000 (95% CI: 980.86–2089.48), respectively (Table 1). For women aged 50--54 years, the ASPR, ASIR, and ASDR were also highest in low-SDI regions, with values of 9122.8 per 100,000 (95% CI: 7616.85–10858.64), 10181.45 per 100,000 (95% CI: 8014.37–12900.12), and 1554.63 per 100,000 (95% CI: 1026.89–2155.23), respectively (Table 2).

From 1990--2021, the ASPR, ASIR, and DALYs for menopausal women in both age groups increased in high-SDI regions (Table 1, Table 2). Notably, among the five SDI regions, only high-SDI regions presented positive EAPCs in ASPR, ASIR, and DALYs (Table 1, Table 2).

### Regional trends

The prevalence of DD among menopausal women has shown significant region al disparities. In 1990, the highest ASPR for menopausal women aged 45--49 years was observed in Central Sub-Saharan Africa, at 10,347.34 per 100,000 (9 5% CI: 8,158.35--12,876.98)(Table 1 Figure 1). The highest ASIR was also in Central Sub-Saharan Africa, at 11,675.26 per 100,000 (95% CI: 8,910.12–15,19 7.39)(Table 1 Figure 2), and the ASDR was similar in Central Sub-Saharan Africa, at 1,786.83 per 100,000 (95% CI: 1,131.14–2,564.48)(Table 1 Figure 3).

**Fig. 1.**
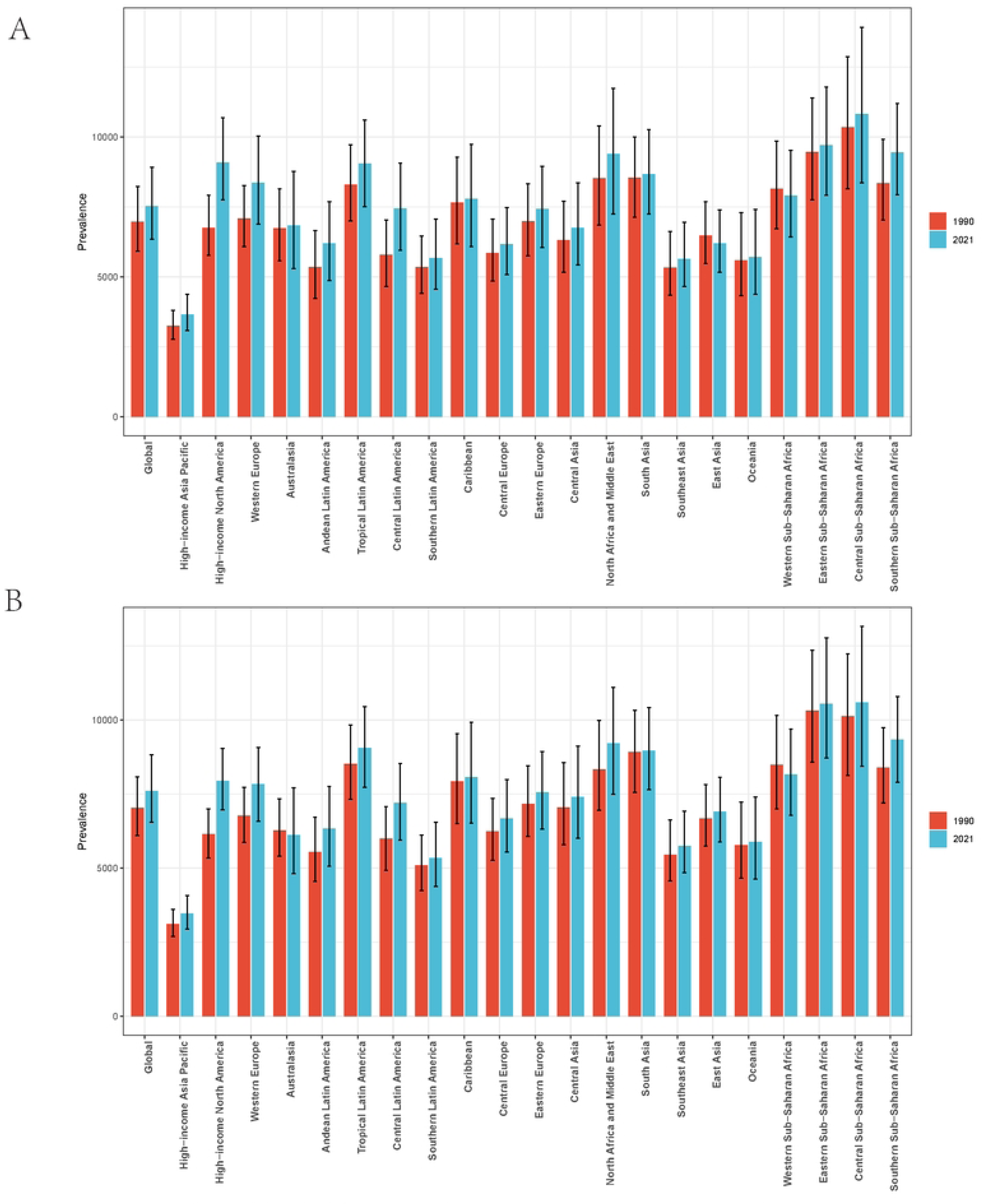
The ASPR of depression among menopausal women in two age groups (A: 45-49 years, B: 50-54 years) across 21 GBD regions.

**Fig. 2.**
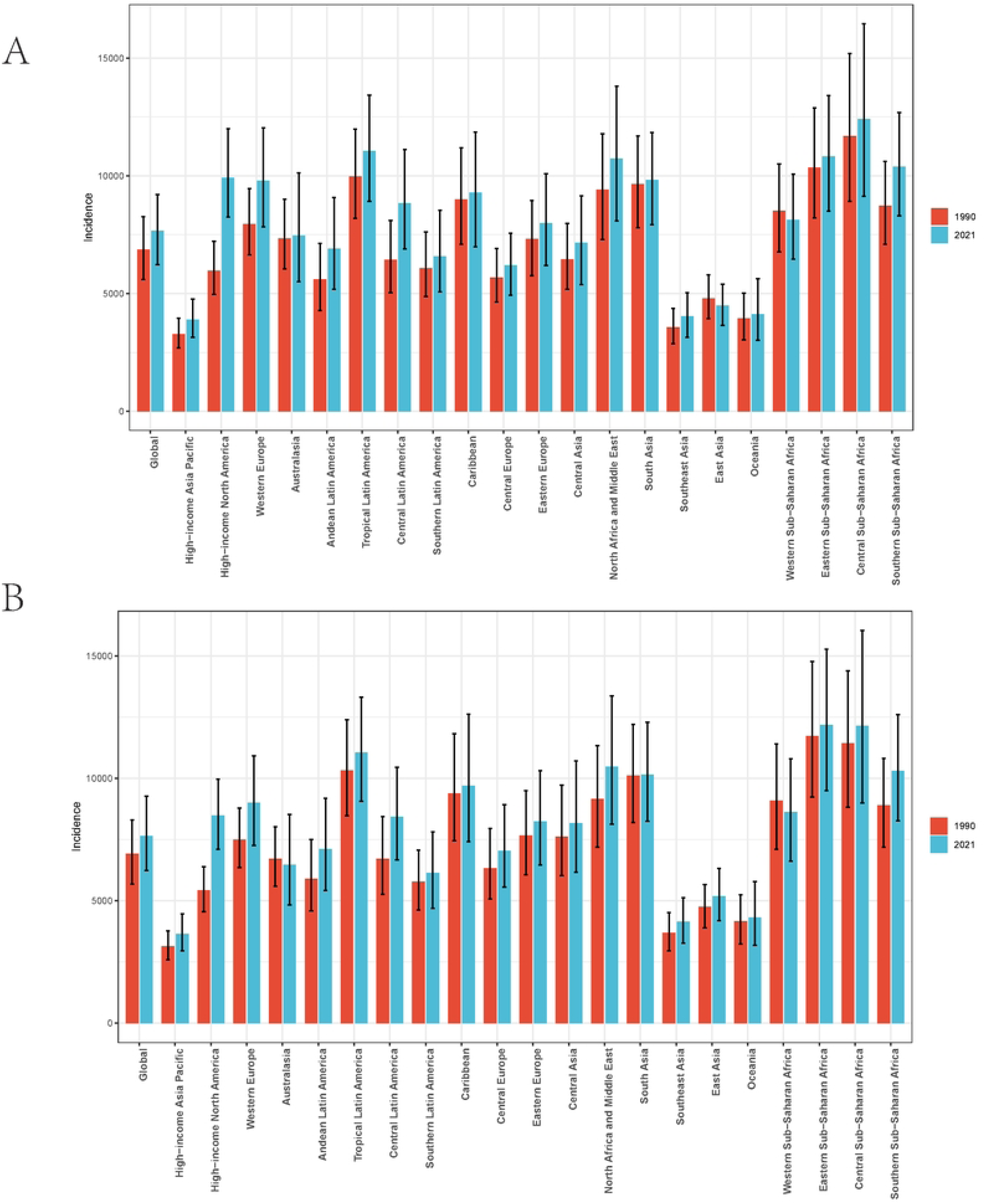
The ASIR of depression among menopausal women in two age groups (A: 45-49 years, B: 50-54 years) across 21 GBD regions.

**Fig. 3.**
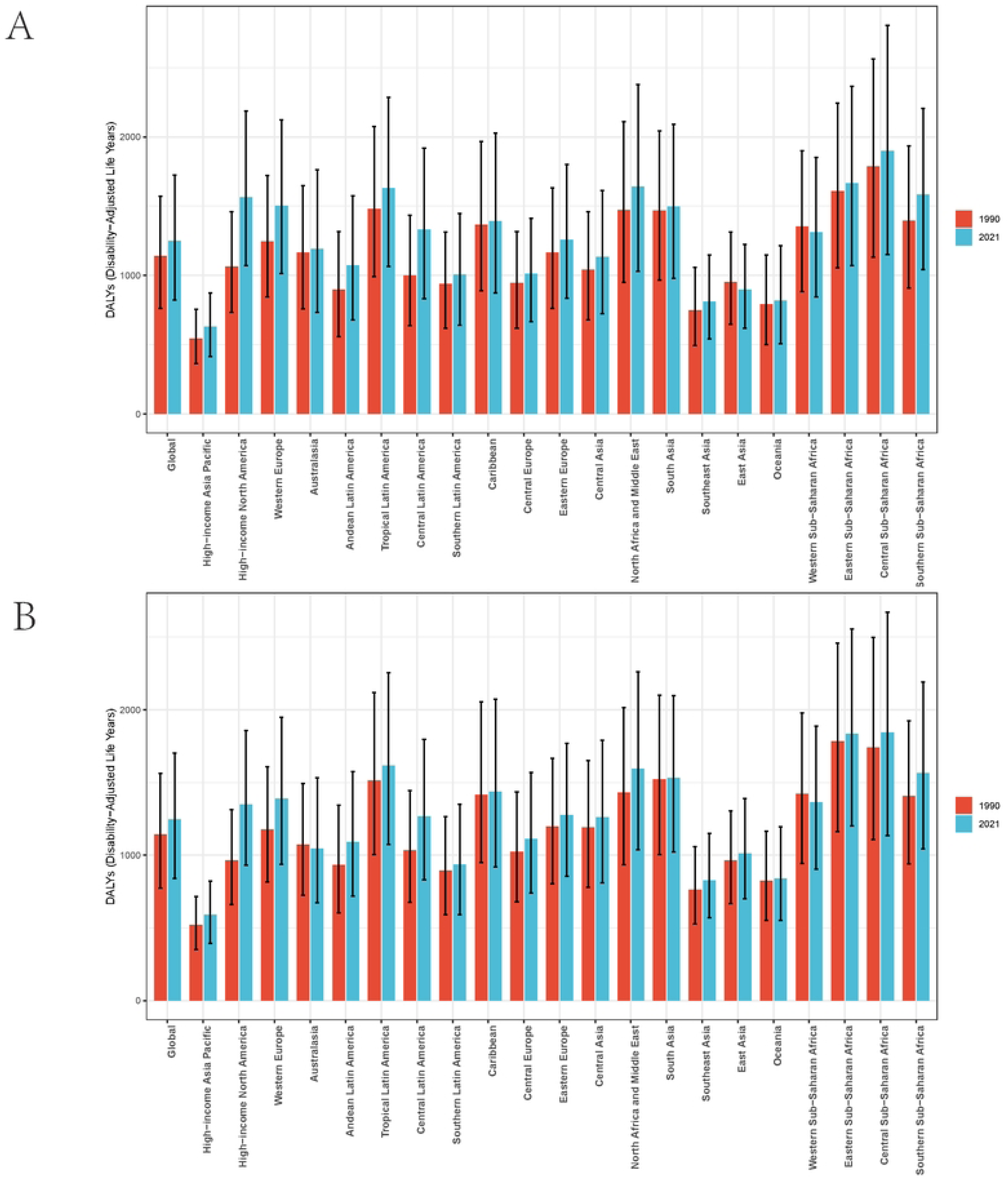
The ASDR of depression among menopausal women in two age groups (A: 45-49 years, B: 50-54 years) across 21 GBD regions.

By 2021, Central Sub-Saharan Africa continued to have the highest ASPR for menopausal women aged 45--49 years, at 10,822.61 per 100,000 (95% CI: 8,36 2.26–13,926.82)(Table 1 Figure 1). The highest ASIR remained in Central Sub-Saharan Africa, at 12,402.31 per 100,000 (95% CI: 9,133.04–16,457.15)(Table 1 Figure 2), and the highest ASDR was also in Central Sub-Saharan Africa, at 1,897.94 per 100,000 (95% CI: 1,150.52–2,806.27)(Table 1 Figure 3).

Among menopausal women aged 50--54 years in 1990, the highest ASPR was in Eastern Sub-Saharan Africa, at 10,313.13 per 100,000 (95% CI: 8,584.85–12, 357)(Table 2 Figure 1); the highest ASIR was also in Eastern Sub-Saharan Africa, at 11,739.9 per 100,000 (95% CI: 9,239.8–14,782.76)(Table 1 Figure 2); and the highest ASDR was in Eastern Sub-Saharan Africa, at 1,781.36 per 100, 000 (95% CI: 1,162.85–2,457.05)(Table 2 Figure 3). By 2021, the highest ASP R for this age group shifted to Central Sub-Saharan Africa, at 10,584.81 per 1 00,000 (95% CI: 8,440.73–13,152.43)(Table 2 Figure 1). The highest ASIR was in Eastern Sub-Saharan Africa, at 12,180.99 per 100,000 (95% CI: 9,495.4–15, 280.37)(Table 2 Figure 2), and the highest ASDR was in Central Sub-Saharan Africa, at 1,842.84 per 100,000 (95% CI: 1,135.83–2,671.53)(Table 2 Figure 3).

From 1990–2021, the most significant increases in the ASPR among menopausal women aged 45–49 years were observed in Central Latin America (EAPCs: 0.58, 95% CI: 0.45–0.71)(Table 1). The highest increase in the ASIR was in high-income North America (EAPCs: 0.91, 95% CI: 0.58–1.24)(Table 1), and the highest increase in the ASDR was in Central Latin America (EAPCs: 0.66, 95% CI: 0.52–0.80)(Table 1).

For menopausal women aged 50--54 years, the greatest increases in ASPR, AS IR, and ASDR were observed in high-income North America, with EAPCs of 0.36 (95% CI: 0.22–0.50), 0.77 (95% CI: 0.49–1.06), and 0.53 (95% CI: 0.33–0.72), respectively(Table 2)

### National trends

In terms of the number of cases reported in 2021, China, India, and the United States had the highest prevalence of DD among menopausal women in both the 45–49 and the 50–54 age groups. Specifically, for the 45–49 age group, the numbers of cases in China, India, and the United States were 3,368,630.622 (95% CI: 2,813,077.473–4,023,770.826), 3,177,688.124 (95% CI: 2,672,521.194–3,733,085.567), and 930,158.9303 (95% CI: 797,852.4047–1,089,847.985), respectively . For the 50–54 age group, the numbers of cases in China, India, and the United States were 4,128,403.773 (95% CI: 3,527,029.032–4,821,949.225), 2, 864,409.366 (95% CI: 2,460,481.289–3,296,334.316), and 864,875.2831 (95% CI: 755,640.5816–983,658.5592), respectively(Supplementary Table 1) .

With respect to the ASPR, in 2021, the highest ASPR for menopausal women aged 45--49 years was observed in Gambia, at 13,709.03 per 100,000 (95% CI: 10,219.10--17,992.10) (Supplementary Table 1,Figure 4). Similarly, for women aged 50--54 years, the highest ASPR was also in Gambia, at 14,601.98 per 100,000 (95% CI: 11,069.54– 19,074.11) (Supplementary Table 1,Figure 4).

**Fig. 4.**
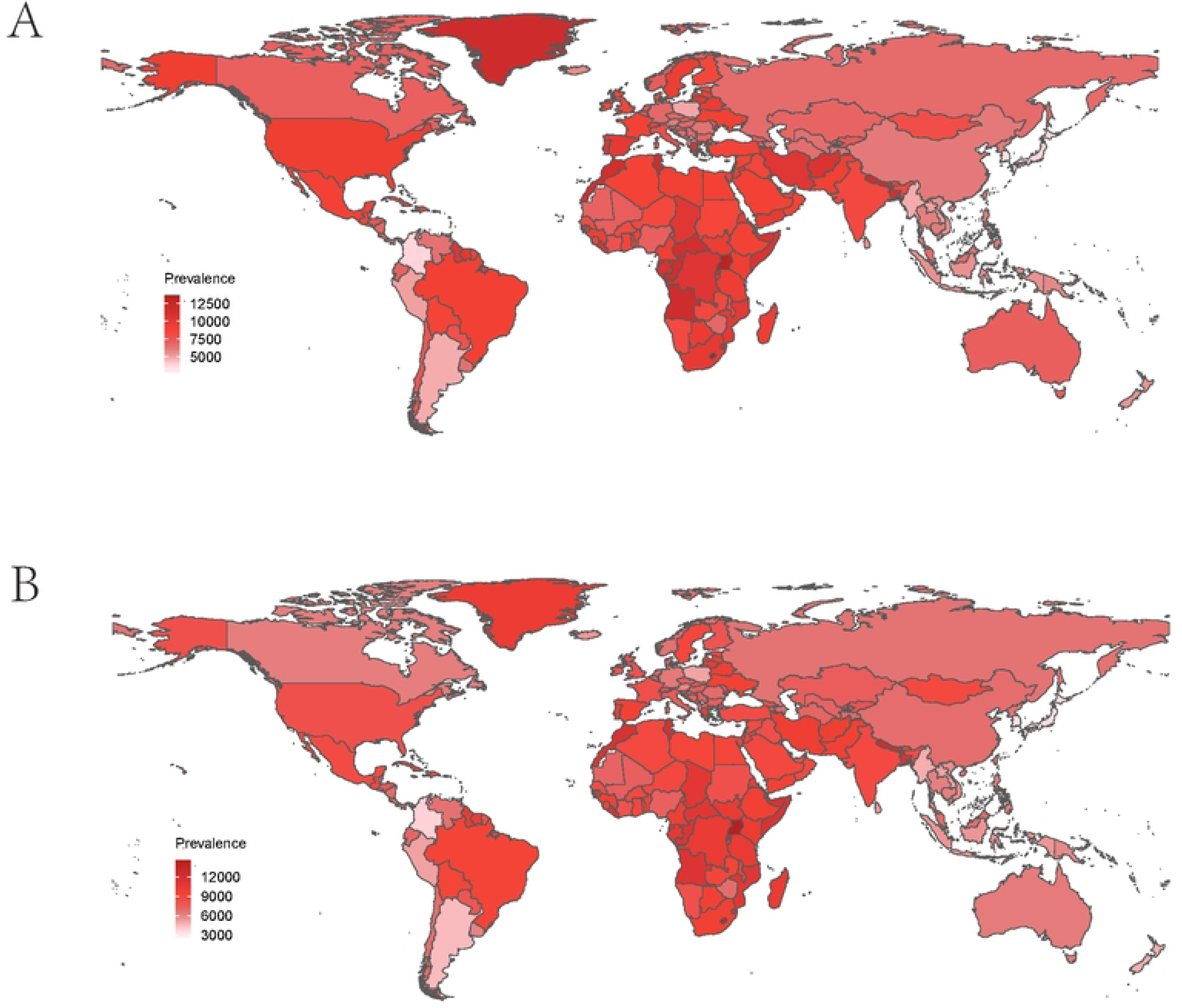
ASPR of depression in menopausal women in two age groups in 204 countries worldwide in 2021. (A 45-49 age group, B 50-54 age group)

In terms of the ASIR, in 2021, Gambia had the highest ASIR for menopausal women aged 45--49 years, at 16,678.27 per 100,000 (95% CI: 11,714.61–23,023.36) (Supplementary Table 1,Figure 5). For the 50–54 year age group, the highest ASIR was again in Gambia, at 18,021.65 per 100,000 (95% CI: 12,768.47–24,855.01) (Supplementary Table 1,Figure 5).

**Fig. 5.**
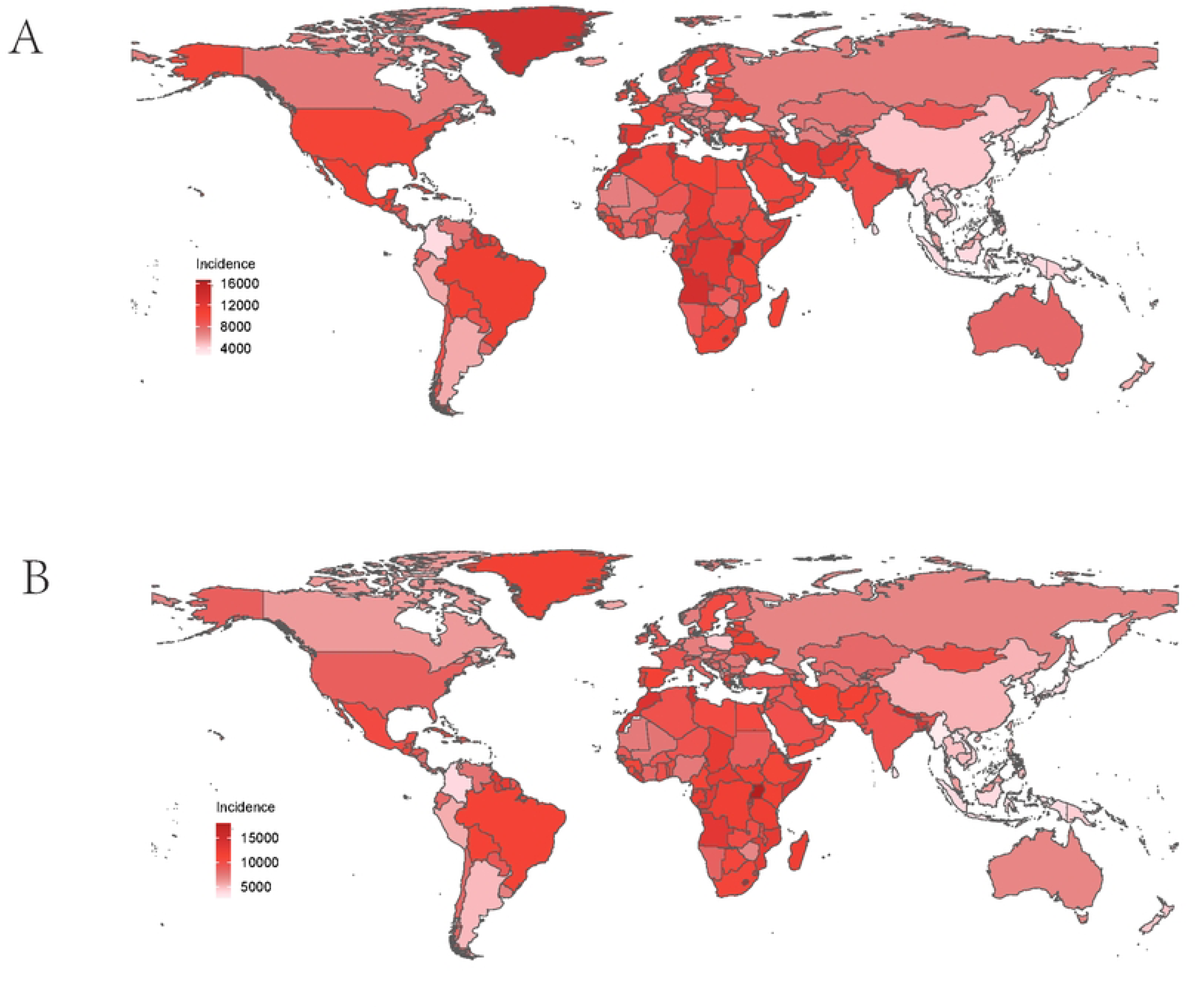
ASPR of depression in menopausal women in two age groups in 204 countries worldwide in 2021. (A 45-49 age group, B 50-54 age group)

For the ASDR, in 2021, Gambia had the highest ASDR for both age groups. For the 45–49 age group, the ASDR was 2,483.98 per 100,000 (95% CI: 1,466.72–3,786.81), and for the 50–54 age group, it was 2,652.75 per 100,000 (95% CI: 1,594.07–4,005.01) (Supplementary Table 1,Figure 6).

**Fig. 6.**
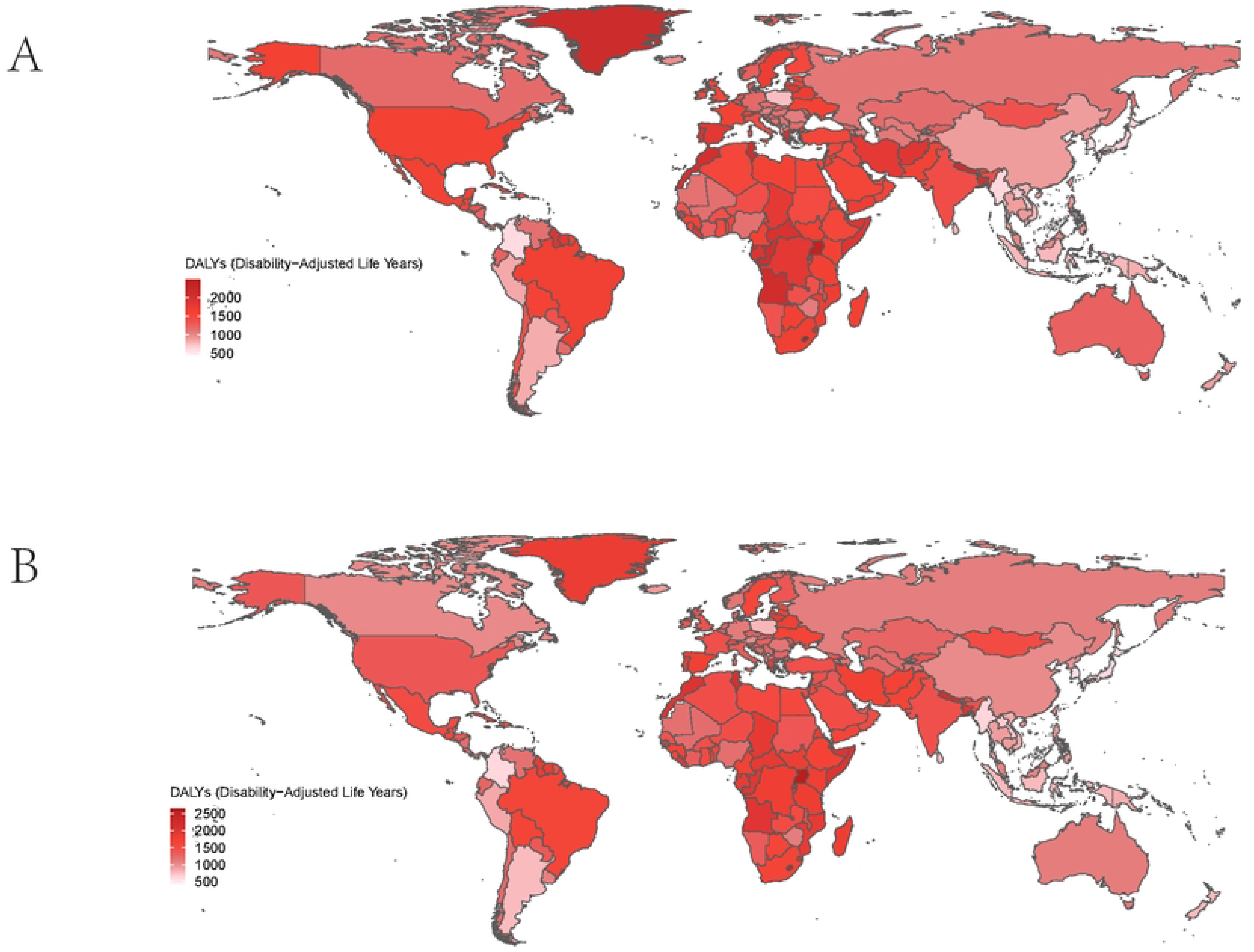
ASDR of depression in menopausal women in two age groups in 204 countries worldwide in 2021. (A 45-49 age group, B 50-54 age group)

## Discussion

Globally, DD among menopausal women has garnered substantial attention in the field of psychiatry but remains inadequately addressed in public health and epidemiological research. Our analysis revealed a steady increase in the burden of DD among menopausal women over the past three decades, as evidenced by the significant increase in prevalence, incidence, and disability-adjusted life years (DALYs). Specifically, from 1990--2021, the ASPR, ASIR, and DALYs for DD among menopausal women aged 45--49 years increased by approximately 123.68%, 130.99%, and 126.46%, respectively. For women aged 50--54 years, these metrics increased by approximately 130.07%, 130.99%, and 126.46%, respectively. In addition to these substantial increases, the burden of DD among menopausal women aged 45--54 years also exhibited geographical imbalances, sociodemographic disparities, and age-structural heterogeneity.

Globally, the ASPR, ASIR, and ASDR of DD among menopausal women in both the 45–49 and 50–54 age groups have significantly increased over the past three decades. However, the EAPCs showed a declining trend in the 45–49 years age group and no significant trend in the 50–54 years ag e group. This pattern indicates that while the burden of DD among menopausal women aged 45--54 years continues to rise, global health efforts have mitigated the burden to some extent. The lack of positive EAPCs in both age groups may reflect the effectiveness of global public health interventions aimed at alleviating depression. International organizations and national governments may have contributed to this reduction through enhanced mental health services, public awareness campaigns, and early intervention programs[15]. Compared with other age groups, the prevalence of DD is greater among menopausal women. Many epidemiological studies rely on self-report questionnaires to explore depressive symptoms in menopausal women, which are less accurate than strict diagnostic procedures for depression and often yield inconsistent results. The prevalence of depression among menopausal women is estimated to be 10.7% in Australia, 36.3% in China, and 42.47% in India[16]. The epidemiological trends and etiological mechanisms of depression in this age group also differ from those in other age groups. Recent studies have shown that depression in menopausal women is closely related to inflammatory mechanisms, sleep quality, hormonal changes, emotional regulation, and social psychology factors [17, 18]. Current treatments for menopausal depression primarily include pharmacotherapy, psychotherapy, traditional Chinese medicine, and lifestyle modifications. Research indicates that while various treatments can improve symptoms, the effectiveness of single treatment modalities is often limited, and a comprehensive treatment plan is usually needed[19, 20]. Additionally, treatment outcomes vary among individuals: some patients may respond well to pharmacotherapy or psychotherapy, whereas others may show no response or poor response. Emerging evidence suggests that hormone replacement therapy, especially during the menopausal period, can effectively prevent postmenopausal depression[21]. Estrogen may have a protective effect on the pathogenesis of depression, and its decline may increase the risk of developing depression[5].

The burden of DD among menopausal women aged 45--49 years is lower than that among women aged 50--54 years. In 2021, the ASPR for the 45–49 age group was 7,529.49 per 100,000 (95% CI: 6,347.01–8,921.84), which was lower than the 7,605.67 per 100,000 (95% CI: 6,552.29–8,818.27) observed in the 50–54 age group. Over 31 years, while the ASPR in the 45–49 years age group showed a declining trend, the trend in the 50–54 years age group was not significant. This discrepancy is related to the different social and physiological challenges faced by menopausal women at different stages[22]. Women in the menopausal period experience gradual ovarian dysfunction and declining estrogen levels[23], which further decrease during the postmenopausal period (ages 50--54)[24]. This results in more pronounced psychological symptoms (such as anxiety and low self-esteem) and somatic symptoms (such as fatigue and insomnia) in postmenopausal women than in those in the 45--49 years[25]. Hormonal changes also increase the risk of cardiovascular diseases and osteoporosis[9]. In addition to hormonal effects, postmenopausal women (ages 50--54) must adapt to changes in their bodies and appearance[26], face the reality of aging, and may develop concerns about health and quality of life[27]. The social roles of postmenopausal women may also shift, such as retirement or becoming empty nesters[28]. These changes can lead to a reevaluation of self-worth and an increased demand for social participation, which in turn exacerbates the prevalence of depression[29].

The burden of DD among perimenopausal women is closely related to the SDI. Specifically, low-SDI regions bear a heavier disease burden, whereas high-SDI regions experience a faster increase in disease burden. The causes of DD in low- and high-SDI regions are significantly different. The heavier burden in low-SDI regions may be related to disparities in medical resources and health care coverage[6]. In low-SDI regions, a vicious cycle exists between poverty and depression: poverty leads to worries about life, poor physical conditions, adverse early-life conditions, social violence, high crime rates, and low social status, all of which often trigger depression. This cycle may also be related to underdeveloped labor forces, children and adolescents, preferences and beliefs, and economic decision-making, while depression further exacerbates poverty [30]. An increasing number of studies have documented the economic, social, and cultural factors behind these differences. For example, in low-income countries in East Africa, such as Gambia, poor eco nomic conditions may affect access to medical resources[31], and women are particularly susceptible to cycles of poverty, low status, and medical inequality[32, 33]. In contrast, high-SDI regions are the fastest-growing areas in terms of the ASIR, ASPR, and DALYs among the five SDI regions. With significant improvements in living standards, residents may experience anxiety and depression due to excessive rumination. For example, the burden of depression is greater in high-SDI countries such as the United States, which may be related to lifestyle factors, income inequality, personal debt, and excessive rumination driven by high socioeconomic status[34, 35], with personal debt having a stronger correlation with the high burden of DD[36]. Additionally, sedentary lifestyles, poor dietary habits, and a lack of physical exercise are more common in affluent societies and can also have negative impacts on mental health[37].

Globally, the burden of DD among menopausal women has significantly increased over the past three decades, with clear trend differences across regions and age groups. Addressing the high prevalence of DD among menopausal women is profoundly important to women’s health. First, strengthening family care is essential, as it plays a crucial role in improving the mental health of postmenopausal women. Good family care can significantly alleviate depressive symptoms by providing emotional support, reducing negative emotions, and enhancing psychological security [18]. Second, enhancing public awareness and screening for depression is vital. Early intervention can significantly increase the disease burden, and optimizing screening procedures and increasing public awareness of depression can effectively improve patient outcomes and reduce the burden on the public health system[10, 38]. Third, national and government policies that alleviate stress in menopausal women are necessary. Stress is an important risk factor for menopausal depression, and stress management and psychological support can help relieve stress during this life stage[39, 40].

In summary, the burden of DD among menopausal women aged 45--54 years continues to increase globally, but the growth rate has been somewhat controlled by efforts from the global health system. The epidemiological trends of DD among menopausal women in the 45–49- and 50–54-year-old age groups differ, with the 45–49-year-old age group having a lower disease burden. The etiological factors of DD among menopausal women vary by age group. In addition to age differences, geographical disparities are also important factors affecting the burden of depression. Low-SDI regions bear a heavier disease burden due to medical conditions, social status, and wealth disparities, whereas high-SDI regions face faster rates of DD incidence and prevalence due to higher diagnostic levels and increased life stress from rapid social development in recent decades.

Although this study utilized rich data from the GBD study, it still has several limitations. GBD data rely on disease surveillance systems and reporting data from various countries, and differences in data quality and completeness may affect the accuracy of the results. Some countries may underreport cases due to differences in medical or diagnostic standards, thereby underestimating the actual disease burden. Additionally, this study did not delve into the risk factors for DD among menopausal women. For example, cultural differences across regions and countries can also influence the occurrence of depressive symptoms, which may interfere with the results.

## Conclusion

This study analyzed the burden of DD among menopausal women aged 45--54 years globally from 1990--2021, revealing trend changes at the global, national, and regional levels. The results indicate that although the burden of DD among menopausal women has significantly increased and has continued to rise steadily over the past three decades, the rate of increase in incidence has been mitigated by efforts from the global health system. Additionally, the burden and etiological factors of DD among menopausal women vary by age group, sociodemographic level, and region.

## Data Availability

All relevant data are within the manuscript and its Supporting Information files.

## Declarations

### Ethics approval and consent to participate

Not applicable.

### Consent for publication

All authors have approved the publication.

### Availability of data and materials

Please direct all data requests to the corresponding author.

### Competing interests

The authors declare that they have no conflicts of interest related to this article.

### Funding

This work was supported by the Scientific Research Project of the Science and Technology Bureau of Nanchong (No. 23JCYJPT0014).

### Authors’ contributions

Y. S.P. was responsible for the initial design of the study, performed the data analysis using the GBD database, and drafted the initial manuscript. Y.F.T. and Y.L.H. contributed to the software development for data processing and validation of the GBD data used in the study, as well as provided critical feedback on the manuscript.

S.L. conducted additional research to support the methodology. W.Z. reviewed the manuscript for content. All authors reviewed the manuscript.

## Acknowledgements

Thanks to the Institute for Health Metrics and Evaluation for the data of the GBD database. We thank Jinding Statistics for kindly providing statistical support.

**Graphical abstract**

Supplementary Tab. 1. The ASPR, ASIR and ASDR of menopausal women in 204 countries and regions from 1990 to 2021.

Supplementary Tab. 2. The number of ASPR, ASIR and ASDR among menopausal women in some countries from 1990 to 2021.

Supplementary Fig. 1. ASPR of depression inmenopausal women in two age groups in 204 countries worldwide in 1990. (A 45-49 age group, B 50-54 age group)

Supplementary Fig. 2. ASIR of depression in menopausal women in two age groups in 204 countries worldwide in 1990. (A 45-49 age group, B 50-54 age group)

Supplementary Fig. 3. ASDR of depression in menopausal women in two age groups in 204 countries worldwide in 1990. (A 45-49 age group, B 50-54 age group)

Supplementary Fig. 4. Changes in depression prevalence among menopausal women aged 45–49 and 50–54 years in 204 countries and territories. (A. The age group of 45 to 49 years old. B. The age group of 50 to 54 years old.)

Supplementary Fig. 5. Changes in depression incidence among menopausal women aged 45–49 and 50–54 years in 204 countries and territories. (A. The age group of 45 to 49 years old. B. The age group of 50 to 54 years old.)

Supplementary Fig. 6. Changes in depression DALYs among menopausal women aged 45–49 and 50–54 years in 204 countries and territories. (A. The age group of 45 to 49 years old. B. The age group of 50 to 54 years old.)

## Notes

### Competing Interest Statement

The authors have declared no competing interest.

### Funding Statement

Yes

